# Spatio-temporal dynamics of leptospirosis in Brazil between 2010 and 2023: identifying high-risk regions and gender-specific patterns

**DOI:** 10.1101/2025.05.24.25328285

**Authors:** Xiang Chen, Paula Moraga

## Abstract

**Background:** Leptospirosis is a globally neglected zoonotic disease with heightened transmission in tropical regions. Brazil bears a disproportionate burden within Latin America, driven primarily by socio-economic vulnerability, urban infrastructure deficiencies, occupational exposure and hydrological factors such as rainfall and flooding. Despite its importance, the spatio-temporal patterns of leptospirosis across Brazil’s diverse regions and populations remain poorly understood. This study aims to examine the spatial distribution, temporal evolution, and gender-specific patterns of leptospirosis risk in Brazil from 2010 to 2023.

**Methods:** We conducted a national-level retrospective analysis using confirmed leptospirosis case and population data aggregated at the microregional level (*n* = 558) from Brazil’s SINAN and DATASUS systems. Descriptive statistics were used to assess overall trends. Spatio-temporal clusters were identified using spatio-temporal scan statistics with a discrete Poisson model. Bayesian spatio-temporal models were then used to estimate annual relative risks (*RR*) and 95% credible intervals across microregions for total, male, and female populations. An interactive dashboard was developed to facilitate result dissemination.

**Results:** A total of 48,190 cases were reported between 2010 and 2023, with males accounting for 80.1%. Significant interannual variation was observed, with peaks in 2011 and 2018 and a sharp decline during the COVID-19 pandemic. Cluster analysis revealed 30 high-risk areas, primarily located in the Acre state, and southern Brazil. *RR* estimates confirmed persistent endemicity in these regions, with some microregions exceeding *RR >* 20 in multiple years. Gender-specific analyses revealed distinct patterns with several microregions demonstrating diverging trends between genders. Temporal and geographic *RR* variation was visualized through interactive maps and trend plots.

**Interpretation:** This study reveals substantial spatial heterogeneity and gender disparities in leptospirosis risk across Brazil. Persistent high-risk areas, particularly in the Amazon Basin and southern states, highlight the need for long-term, location-specific public health interventions. The divergence in male and female risk trajectories underscores the importance of gender-disaggregated surveillance and targeted prevention strategies. Integrating high-resolution spatio-temporal modeling with interactive tools can support more equitable and data-driven disease control in Brazil and similar endemic settings.

## 1 Introduction

Leptospirosis, a globally emerging zoonotic disease caused by spirochete bacteria of the genus *Leptospira*, poses a significant public health challenge, particularly in tropical and subtropical regions. The disease is transmitted primarily through direct or indirect contact with water, soil, or surfaces contaminated by the urine of infected animals. Environmental factors such as heavy rainfall, flooding, and inadequate sanitation exacerbate its transmission, especially in regions with limited resources and poor infrastructure. Globally, leptospirosis is among the leading zoonotic causes of morbidity and mortality, with an estimated 1.03 million cases and nearly 59,000 deaths annually [1]. However, the true burden is likely underreported due to frequent misdiagnosis, limited clinical awareness, and insufficient surveillance systems in many affected areas [2].

Since the discovery of *Leptospira* in Japanese mine workers over a century ago, leptospirosis has been considered a “neglected tropical disease” [3]. Despite its global distribution and recognition as an emerging infectious disease, it continues to disproportionately affect impoverished populations in low- and middle-income countries [4]. In rural areas, subsistence farmers, rice and sugarcane workers, and other outdoor laborers face heightened risk due to occupational exposure. In urban settings, particularly in densely populated slums, poor sanitation, accumulated waste, and rodent infestations create ideal conditions for sustained transmission [5, 6]. Recreational activities involving contact with contaminated water and occupations such as animal husbandry, sewage work, and farming further contribute to the risk profile.

Brazil, with its vast geographical and climatic diversity, bears a significant portion of the leptospirosis burden in Latin America. The region reports approximately 10,000 human cases annually, with Brazil alone accounting for 40% of these [7]. This disproportionate burden is linked to Brazil’s tropical and subtropical ecosystems, frequent flood events, and deep-rooted socio-economic disparities. Following Brazil, Peru (23.6%), Colombia (8.8%), and Ecuador (7.2%) report the highest number of cases [7], reflecting the broader vulnerability of Latin America to leptospirosis. The highest morbidity rates are observed in resource-limited settings, where surveillance and control measures remain insufficient.

Climatic events such as hurricanes and floods frequently trigger outbreaks, highlighting the role of environmental variability in leptospirosis transmission [8, 9, 10]. In Latin America, the intersection of climatic hazards and socio-economic vulnerabilities has intensified the disease’s impact. Brazil’s unique combination of environmental, demographic, and infrastructural factors necessitates a comprehensive investigation into the spatio-temporal dynamics of leptospirosis. Such analyses are essential to inform tailored prevention and control strategies, which remain underdeveloped in the existing literature [2].

Despite growing recognition of leptospirosis as a significant public health threat, its spatial and temporal determinants remain undercharacterized, particularly at subnational levels [11, 12, 13, 14]. To address these knowledge gaps, this study investigates the spatio-temporal dynamics of leptospirosis in Brazil from 2010 to 2023. Using national surveillance data and advanced spatial and temporal modeling techniques, we quantify disease burden, identify persistent high-risk areas, and evaluate gender-specific risk trajectories across 558 microregions. By generating high-resolution risk estimates and interactive visualizations, this study supports more precise, equitable, and data-driven strategies for leptospirosis control in Brazil and comparable endemic settings.

## 2 Material and methods

In this study, we examined the spatio-temporal dynamics of leptospirosis in Brazil from 2010 to 2023 at the microregional level. By leveraging annual leptospirosis case data and population estimates across 558 microregions, we conducted a comprehensive analysis combining descriptive statistics, spatio-temporal cluster detection, and Bayesian spatial modeling. This multi-method approach allowed us to identify high-risk areas, temporal trends, and gender-specific disparities in disease burden. This analysis provides valuable insights to inform targeted interventions and improve public health strategies in Brazil.

### 2.1 Study area

Brazil spans approximately 8.5 million square kilometers, and remains the most populous country in Latin America with an estimated 212.6 million people as of July 1, 2024, according to the Brazilian Institute of Geography and Statistics (Instituto Brasileiro de Geografia e Estatística, IBGE) [15]. Brazil is characterized by distinct climatic, ecological, and socio-economic profiles that play a crucial role in the dynamics of leptospirosis transmission. The country’s vast population and diverse climatic conditions necessitate a thorough investigation of leptospirosis dynamics to inform targeted interventions.

### 2.2 Data collection

The leptospirosis case data were obtained from the Sistema de Informação de Agravos de Notificação (SINAN, Information System for Notifiable Diseases) [16], Brazil’s national epidemiological surveillance platform. SINAN compiles data on various notifiable diseases, including leptospirosis, through standardized reporting protocols across the country. For this study, we extracted annual counts of confirmed leptospirosis cases between 2010 and 2023 for all 558 microregions. The data were disaggregated by gender, allowing stratified analyses for total, male, and female populations.

Corresponding annual population estimates were obtained from DATASUS (Departamento de Informática do SUS, Department of Information Technology of the Unified Health System) [17]. These estimates, based on data from IBGE, provide microregional-level population counts stratified by gender for the period 2010–2023. The data were used for calculating standardized incidence rates in risk modeling.

All datasets were cleaned, harmonized, and matched spatially using standardized geographic codes provided by IBGE. Microregions without any reported cases were retained in the dataset and assigned zero-case counts to ensure consistency in spatio-temporal analysis.

### 2.3 Descriptive statistical analysis

Initially, descriptive statistical analysis was conducted to provide an overview of lep-tospirosis trends across Brazil. This analysis included bar plots comparing the absolute number of reported cases between males and females, as well as line charts illustrating the annual total number of cases from 2010 to 2023. These visualizations, presented in Figure 1, highlight temporal trends and gender-specific disparities in leptospirosis inci-dence, offering insights into the evolving patterns of disease burden across demographic groups.

**Figure 1:**
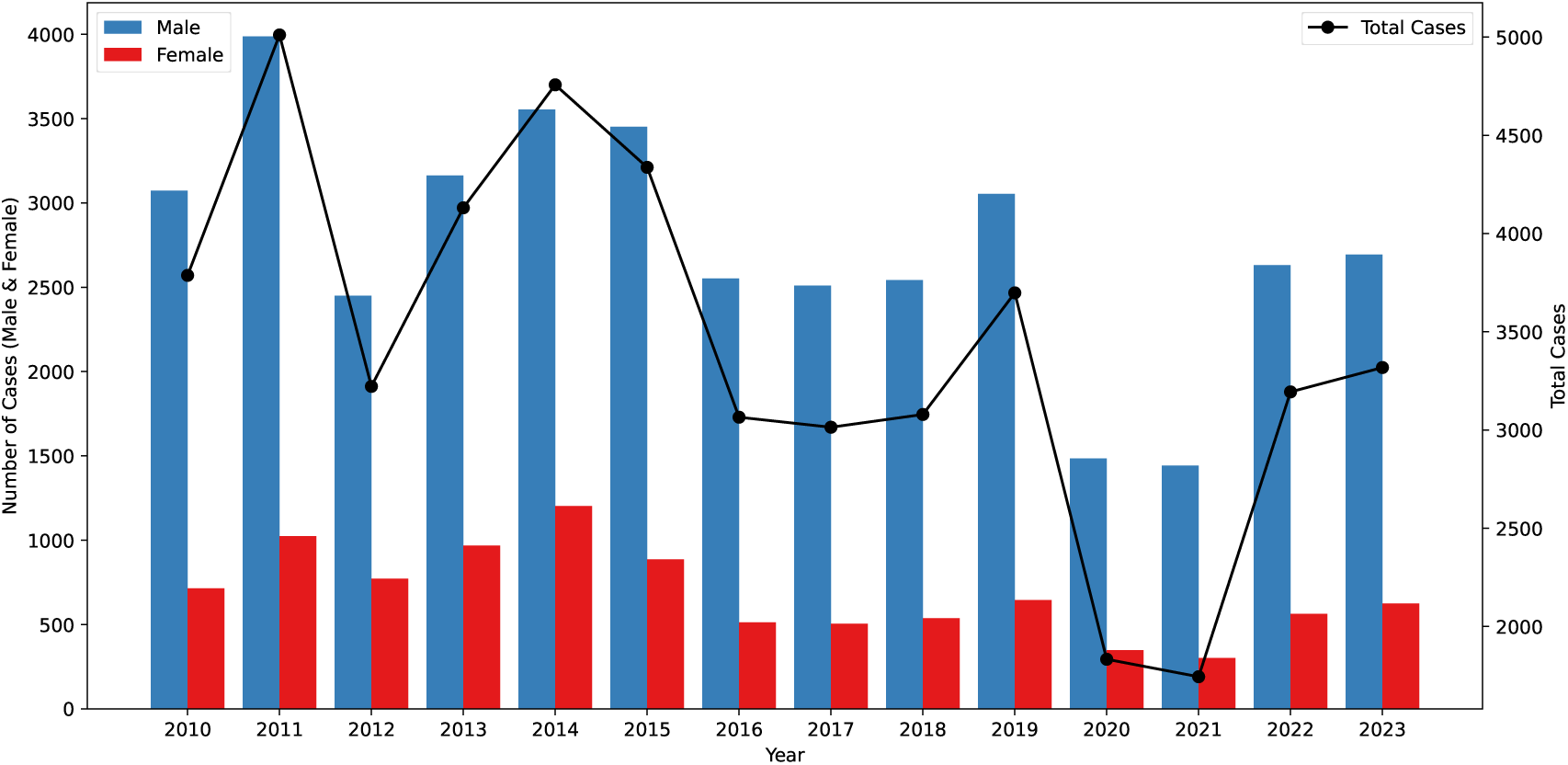
Leptospirosis cases by gender and total in Brazil (2010–2023). Bars represent male (blue) and female (red) case counts (left *y*-axis), while the black line shows total annual cases (right *y*-axis). Note that the left and right axes have different starting points.

### 2.4 Spatio-temporal cluster detection

To identify significant spatio-temporal clusters of leptospirosis in Brazil, we employed spatio-temporal scan statistics via the SaTScan software (version 10.2.5) [18, 19], a widely used tool for detecting spatial, temporal, and space-time disease clusters [20, 21]. SaTScan implements the spatial and spatio-temporal scan statistics, which systemati-cally scan the study area using moving circular or cylinder windows of varying size to detect regions and periods of time with unexpectedly high or low case counts.

The clustering analysis was conducted using the spatio-temporal discrete Poisson probability model, which is appropriate for aggregated count data such as the number of leptospirosis cases per microregion. Under this model, the number of observed cases in each scanning window is compared to the expected number under the null hypothesis of spatial and temporal randomness. A likelihood ratio test is used to evaluate each potential cluster, and statistical significance is assessed through Monte Carlo hypothesis testing with 999 replications.

This method enables the detection of statistically significant spatio-temporal clusters that represent regions and periods with elevated leptospirosis incidence. Identifying such high-risk spatio-temporal clusters is essential for timely public health response and targeted allocation of surveillance and control resources.

### 2.5 Bayesian spatio-temporal modeling

To further estimate the relative risk (*RR*) of leptospirosis across Brazil’s microre-gions while accounting for spatial structure and temporal variability, we implemented a Bayesian spatio-temporal model [22]. Bayesian inference was performed using the Integrated Nested Laplace Approximation (INLA) method [23]. INLA provides a com-putationally efficient alternative to traditional Markov Chain Monte Carlo (MCMC) methods for approximate Bayesian inference in latent Gaussian models, making it es-pecially suitable for large-scale spatial epidemiological analyses.

Given the *i* (*i* = 1*, …, N* = 558) microregions and *t* (*t* = 2010*, …,* 2023) years, let *y_it_* represent the observed number of leptospirosis cases in microregion *i* and year *t*, *P_it_* the population at risk, and *E_it_* the expected number of cases for that region-year, computed via indirect standardization. The expected number of cases was derived by applying the national incidence rate to each microregion’s population.

The number of observed cases *y_it_* was modeled as a Poisson-distributed random variable:

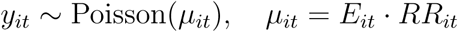

where *µ_it_* is the mean of the Poisson distribution, *E_it_* is the expected number of cases, and *RR_it_* is the relative risk in microregion *i* and year *t*. The relative risk *RR_it_* quantifies whether a region-year exhibits a higher (*RR_it_ >* 1) or lower (*RR_it_ <* 1) leptospirosis risk compared to the national average [24].

The logarithm of the relative risk was modeled as a linear combination of fixed and random effects to capture spatio-temporal variation:

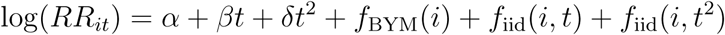

Here, *α* is the overall intercept representing the baseline national risk, and *β* and *δ* are the coefficients for the national-level linear and quadratic temporal time terms, respectively. *f*_BYM_(*i*) denotes the Besag-York-Mollíe (BYM) model [25] for spatially structured and unstructured random effects for microregion *i*, *f*_iid_(*i, t*) represents the unstructured interaction between region and linear temporal term, and *f*_iid_(*i, t*^2^) repre-sents the unstructured interaction between region and quadratic temporal term.

The spatial structure was defined through a neighborhood adjacency matrix, assum-ing microregions that share boundaries are neighbors. The spatially structured compo-nent of the BYM model captures local spatial correlation, while the unstructured term accounts for region-specific heterogeneity. All hyperparameters were assigned vague, non-informative priors, allowing the observed data to primarily drive the inference.

The model was fitted separately for the total population, males, and females to enable gender-disaggregated risk estimation. Posterior summaries, including the pos-terior mean and 95% credible intervals of the relative risk *RR_it_*, were obtained for each microregion-year combination. All models were implemented using the R-INLA package [23], enabling efficient estimation over the full spatio-temporal domain.

This modeling framework enhances understanding of the spatial distribution and temporal evolution of leptospirosis risk, providing crucial evidence to support geograph-ically and demographically targeted public health interventions in Brazil.

## 3 Results

### 3.1 Descriptive analysis

The analysis of leptospirosis cases in Brazil from 2010 to 2023 reveals notable trends and interannual variability. A total of 48,190 confirmed cases were reported nationwide over the study period, averaging approximately 3,442 cases per year. Across all years, males consistently accounted for the majority of cases, comprising roughly 80.1% of the total, while females represented 19.9%. This pronounced gender disparity likely reflects differences in occupational and environmental exposures, with males more fre-quently engaged in activities that increase the risk of contact with contaminated water or environments [26].

The national trend demonstrates substantial year-to-year fluctuations. The highest number of cases was recorded in 2011, exceeding 5,000 nationwide—potentially indicat-ing a severe outbreak or environmental drivers such as heavy rainfall or flooding [27, 28]. In contrast, a marked decline occurred in 2020 and 2021, with reported cases falling to 2,132 and 1,744, respectively. This sharp reduction coincided with the COVID-19 pandemic, during which disruptions to mobility and behavior may have reduced ex-posure to leptospirosis risk factors. Following this dip, a moderate rebound in cases was observed after 2021, signaling a return to pre-pandemic patterns of exposure and transmission.

Despite these fluctuations, the proportional distribution of cases by gender remained remarkably stable, highlighting a persistent elevated risk among male populations. Ad-ditional peaks in 2014 and 2019 further underscore the potential influence of environ-mental and climatic variability on annual case burdens. These patterns are illustrated in Figure 1, which presents bar plots comparing male and female case counts and a line chart depicting total annual cases over time.

### 3.2 High risk clusters

To investigate localized excess risk of leptospirosis, we performed a retrospective space-time scan statistic using SaTScan with a discrete Poisson model, yearly time aggregation, and a scanning window set to detect high-rate clusters. This method identified 30 statistically significant space-time clusters across Brazil from 2010 to 2023, each defined by a spatial area and a specific time interval. Figure 2 displays the spatial and temporal distribution of the detected clusters. Clusters provide insights into both persistent endemic zones and temporally concentrated outbreaks.

**Figure 2:**
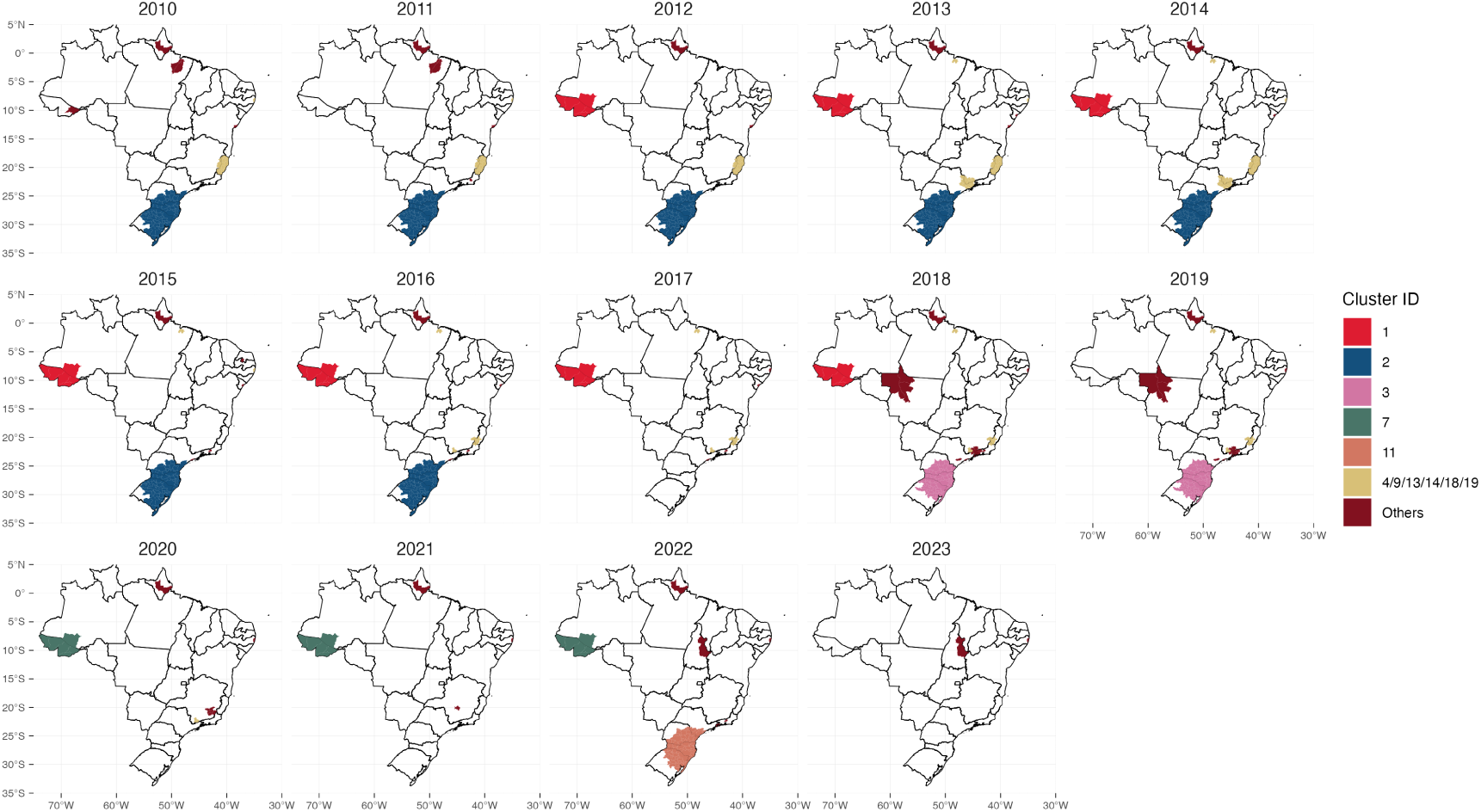
Spatio-temporal distribution of detected clusters across Brazil (2010–2023)

The most intense cluster was found in the Acre state (cluster 1), in the western Amazon. This region exhibited a remarkably elevated *RR* of 37.92 between 2012 and 2018, with 3,554 observed cases compared to only 101 expected. The annual incidence exceeded 115 cases per 100,000 population, marking this area as one of the most hyperendemic leptospirosis zones in the country during the study period. A subsequent space-time cluster was again detected in Acre between 2020 and 2022 (cluster 7), with a similarly elevated *RR* of 8.48, reinforcing the evidence of long-term persistent transmission in this Amazonian frontier.

The southern region of Brazil also emerged as a major area of concern, where mul-tiple large clusters were detected spanning Rio Grande do Sul, Santa Catarina, and Paraná. From 2010 to 2016 (cluster 2), this zone exhibited widespread elevation in risk, with more than 8,400 cases and an *RR* of 3.5, covering over 12 million people. Interestingly, a follow-up cluster re-emerged in the same area during 2018–2019 (cluster 3), and again in 2022 (cluster 11), albeit with slightly lower *RR* values. These patterns suggest a shifting but sustained regional burden, in which high-risk areas evolve tempo-rally yet remain spatially concentrated in southern Brazil. The recurrence of clusters in these states underscores the chronic vulnerability of this region, likely linked to climatic factors such as seasonal flooding, urbanization, and agricultural exposure.

Another noteworthy finding comes from the north, particularly in Beĺem and Ananin-deua in the state of Paŕa. A spatial-temporal cluster spanning 2013 to 2019 showed elevated risk in this densely populated Amazonian area. Despite the lower absolute case counts compared to southern states, the *RR* reached 1.94, with persistent eleva-tion over multiple years. This finding contributes to a growing recognition of neglected leptospirosis risk in northern urban centers, where high rainfall, poor drainage, and peri-urban settlement patterns may foster suitable conditions for transmission.

Additional space-time clusters were also identified along the southeastern coastal regions, including Espírito Santo, Pernambuco, Amapá, São Paulo, and Rio de Janeiro (clusters 4/9/14/18/19). These clusters varied in both size and duration—some were localized and short-lived, such as in Nova Friburgo (RJ), which experienced a sharp outbreak in 2011 with an exceptionally high *RR* exceeding 42, while others persisted across multiple years with moderate but significant risk elevation. These coastal areas, often characterized by dense urban populations, precarious sanitation, and frequent flooding, represent important zones of episodic leptospirosis transmission that may be overlooked in broader national-level analyses. This diversity in cluster duration and spatial size highlights the multifactorial nature of leptospirosis transmission: while some outbreaks are short-lived and acute, others reflect sustained environmental and socio-structural risk.

Notably, while most clusters were spatially discrete in a given time frame, they frequently overlapped geographically across different periods. This temporal fragmentation of clusters in southern Brazil, for example, signals not a one-time outbreak but a persistent endemicity interrupted by transient declines and returns—possibly driven by hydrometeorological factors (e.g., rainfall and flooding), fluctuations in rodent reservoir populations, or variations in public health response and infrastructure coverage.

These high-risk clusters, particularly in the west (Acre) and in the southern states (Rio Grande do Sul, Santa Catarina, and Parańa), and selected northern and eastern regions, offer key geographic targets for intervention, surveillance enhancement, and risk mitigation policies. They also support the findings of the Bayesian spatio-temporal model (Section 3.3), where many of the same microregions exhibited *RR >* 1 across multiple years.

To enhance usability of these results by public health officials, we provide a dynamic web-based viewer, allowing users to interactively explore cluster locations, time spans, and risk intensities: https://www.paulamoraga.com/leptobrazilclusters/. The online map complements static visualizations by enabling temporal comparisons and detailed inspection of microregional patterns, thereby serving as a practical tool for surveillance and preparedness.

### 3.3 Spatio-temporal analysis of leptospirosis risk

The relative risk (*RR*) of leptospirosis infection across 558 microregions in Brazil was assessed yearly from 2010 to 2023. Using a Bayesian spatio-temporal model, *RR* val-ues were computed based on observed leptospirosis cases and population data, with expected cases derived via indirect standardization. The resulting posterior estimates were visualized as annual maps to reveal spatial and temporal patterns in disease risk (Figure 3).

**Figure 3:**
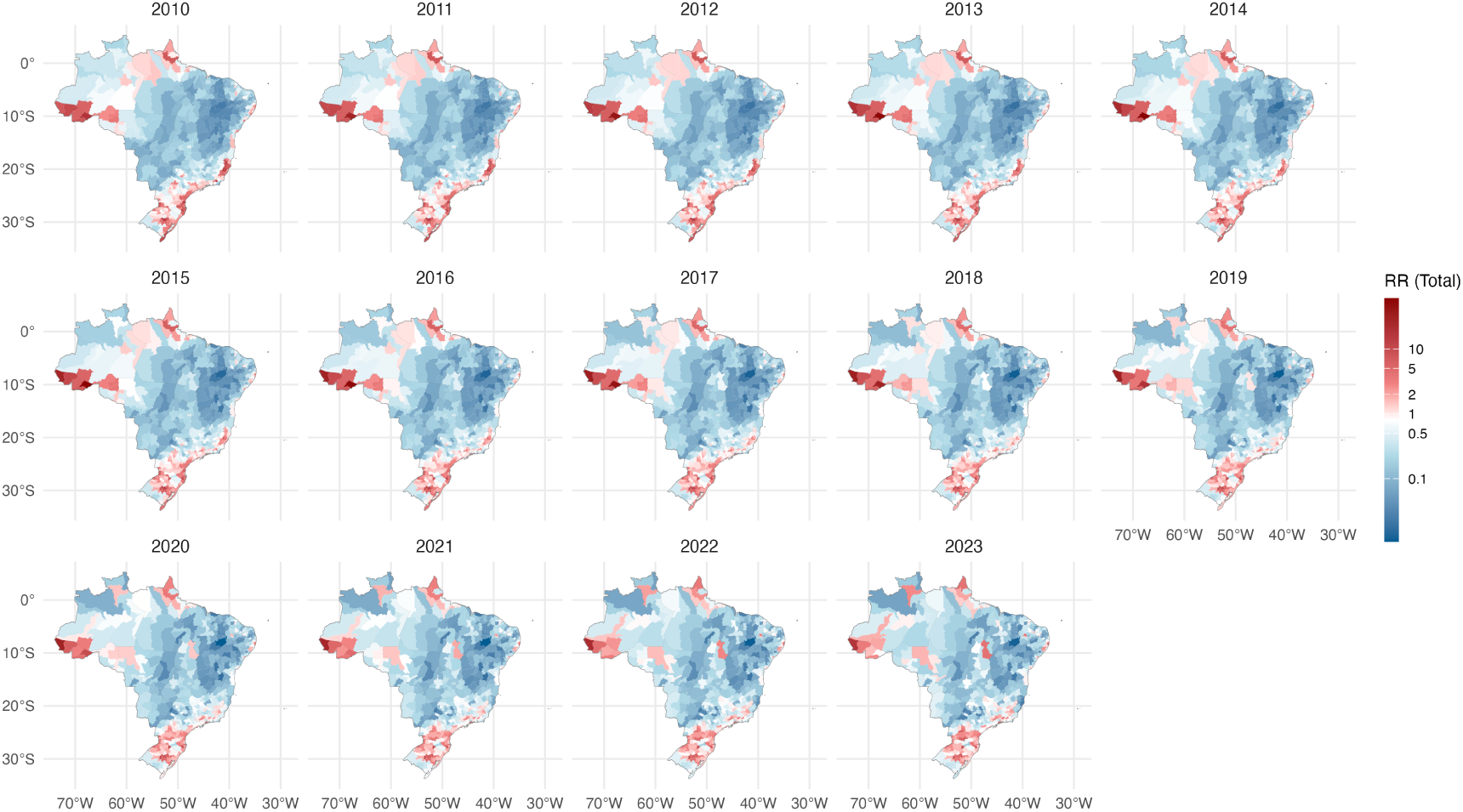
Relative risk (total) of leptospirosis in Brazil (2010–2023). Microregion-level estimates (*RR >* 1 indicates elevated risk)

Throughout the study period, *RR* estimates showed persistent geographic hetero-geneity. Microregions in the southern states—particularly Rio Grande do Sul, Santa Catarina, and Parańa—consistently exhibited elevated risks, aligning with the SaTScan-identified clusters described in Section 3.2. These regions showed *RR* values frequently exceeding 2 and, in some microregions, reaching values above 5. Similar patterns were observed in parts of Acre, Rond^onia, and Mato Grosso, reinforcing their designation as persistent high-risk areas.

Temporal variation was also evident, with risk intensifying in specific periods—especially during years with higher national case counts (e.g., 2011, 2013, and 2018). By con-trast, *RR* values were markedly lower in 2020 and 2021, coinciding with the COVID-19 pandemic, which may have reduced exposure to environmental sources of infection or disrupted disease reporting.

These model-based *RR* estimates corroborate the findings from the purely spatio-temporal cluster analysis, with high-risk zones overlapping substantially with detected clusters. Moreover, the spatio-temporal maps revealed dynamic changes in risk inten-sity and geographic spread, demonstrating that while certain areas maintain persis-tently high risk, others experience episodic surges likely influenced by environmental or socioeconomic factors.

To better characterize persistent high-risk zones, we identified all microregions where *RR >* 1 occurred in at least one year. These are mapped in Figure 4, with color intensity reflecting the number of years each microregion remained above this high-risk threshold. Notably, clusters of long-term elevated *RR* are concentrated in the Amazon Basin and the South, reinforcing regional inequalities in exposure.

**Figure 4:**
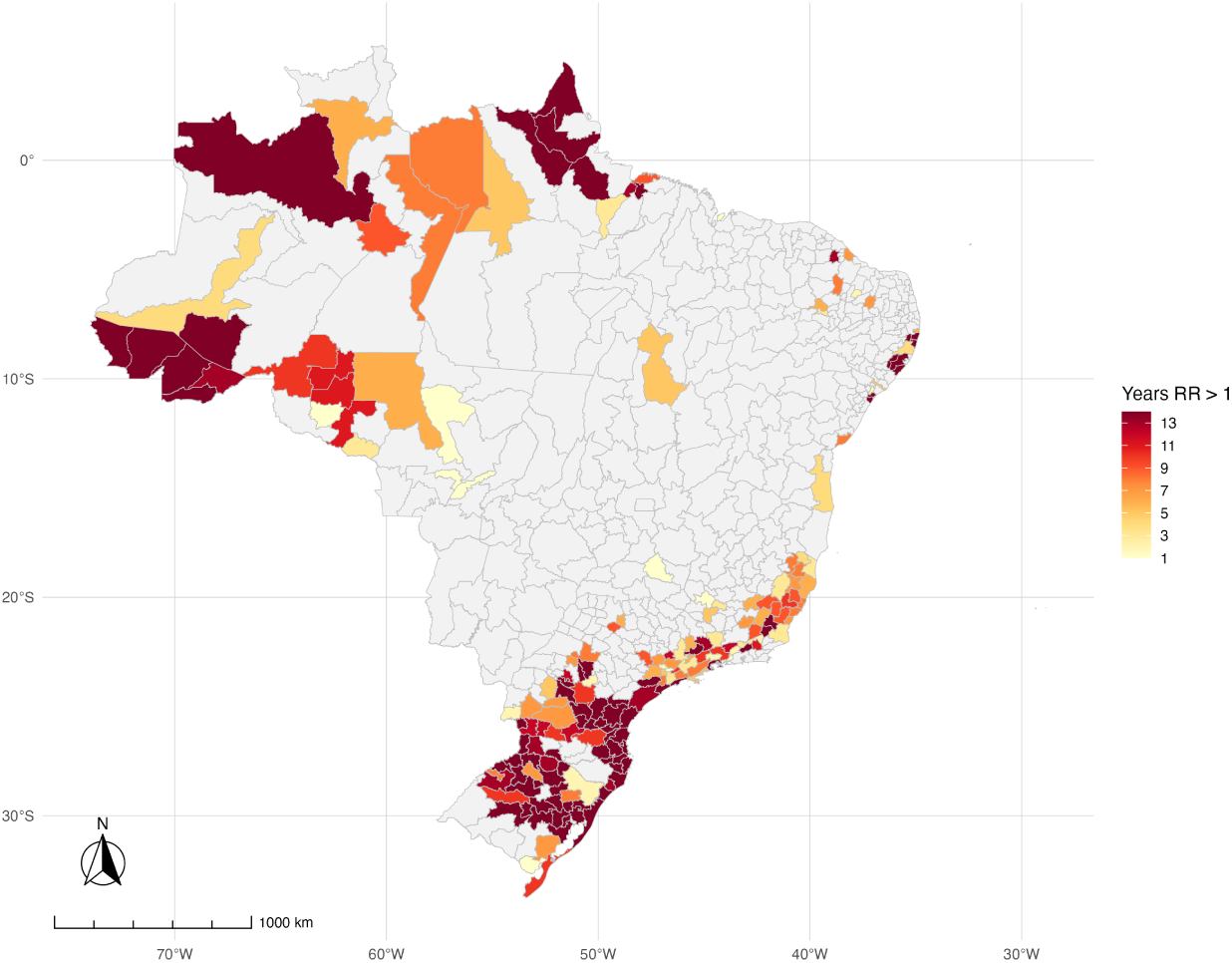
Microregions with Leptospirosis *RR >* 1 (2010–2023). Color intensity reflects the number of years with elevated *RR*.

To further investigate extreme cases, we selected the top 5 microregions with the highest *RR* in any given year from 2010–2023: Cruzeiro Do Sul, Tarauaća, and Rio Branco (Acre state); Afonso Cĺaudio (Espírito Santo state); Fernando de Noronha (Pernambuco state); Nova Friburgo (Rio de Janeiro state); Sananduva, Restinga Seca, and Santa Cruz Do Sul (Rio Grande do Sul state); Tabuleiro (Santa Catarina state). Their *RR* trajectories over time are plotted in Figure 5, with a red dashed reference line at *RR* = 1. The patterns reveal important differences:

**Figure 5:**
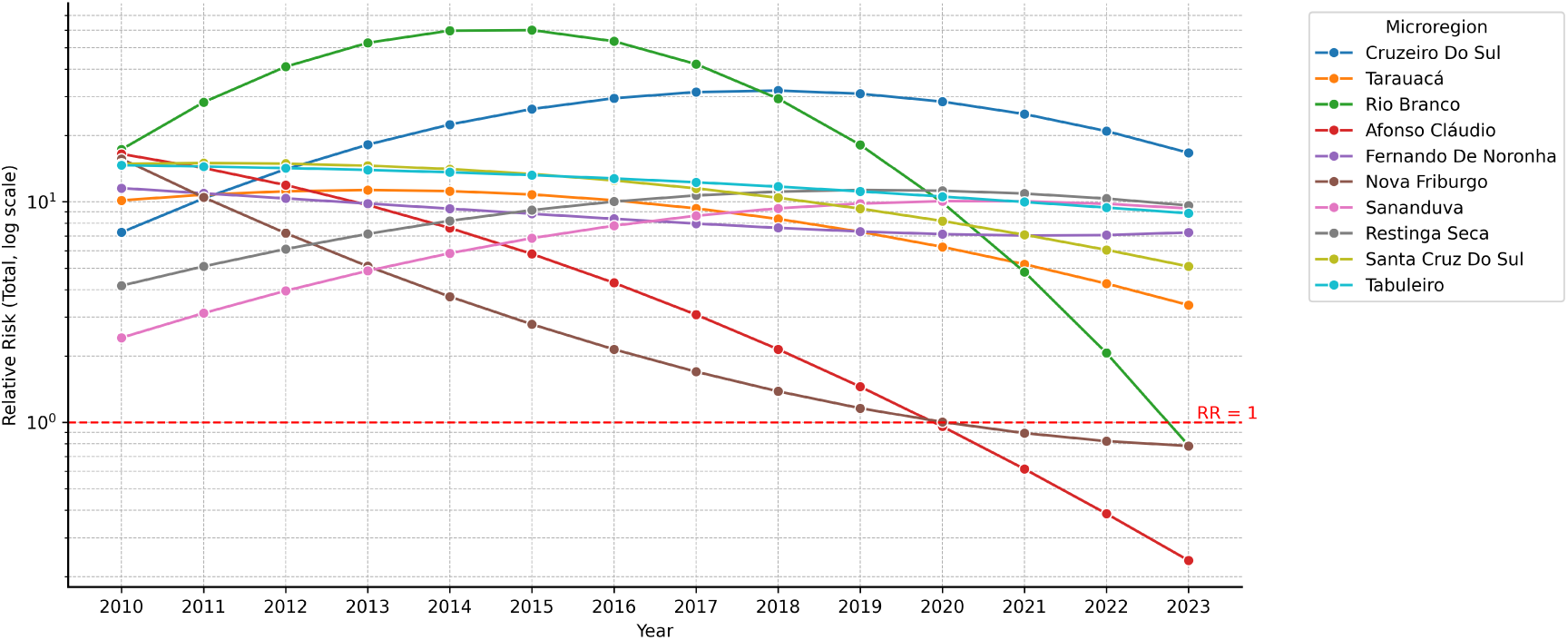
Relative risk (total) over time for top 5 microregions from 2010 to 2023 each year.

Rio Branco experienced a dramatic surge, peaking at *RR* = 59.76 (95% CI: [57.29, 62.31]) in 2014 and *RR* = 60.08 (95% CI: [57.50, 62.74]) in 2015 before dropping sharply, sug-gesting outbreak dynamics superimposed on endemic risk. Cruzeiro do Sul and Ta-rauacá showed long-term elevation with mid-decade peaks, consistent with sustained transmission in remote Amazonian municipalities. In contrast, Afonso Cĺaudio and Nova Friburgo displayed high risk in the early 2010s (e.g., *RR* = 16.51 (95% CI: [13.18, 20.42]) in 2011) followed by steady declines to below 1.0 by 2023—indicating possible impact of control efforts or changing environmental exposures. Sananduva, Restinga Seca, and Santa Cruz Do Sul exhibited stable *RR* values between 5–10 across most of the study period, reflecting entrenched endemicity in flood-prone rural areas of southern Brazil. Tabuleiro showed intermittent peaks, suggesting episodic surges rather than sustained transmission.

These diverse trajectories underscore the value of spatio-temporal modeling: while some regions experience occasional outbreaks, others endure persistent high risk, re-quiring long-term structural interventions rather than reactive response.

### 3.4 Gender-specific patterns

Throughout the 2010 to 2023 timeframe, the spatio-temporal analysis of leptospirosis risk among males and females revealed distinct spatial and temporal heterogeneity (Figures 6 and 7). Although overall patterns mirrored the total population results, gender-specific maps highlighted clear disparities in risk intensity and persistence across microregions.

**Figure 6:**
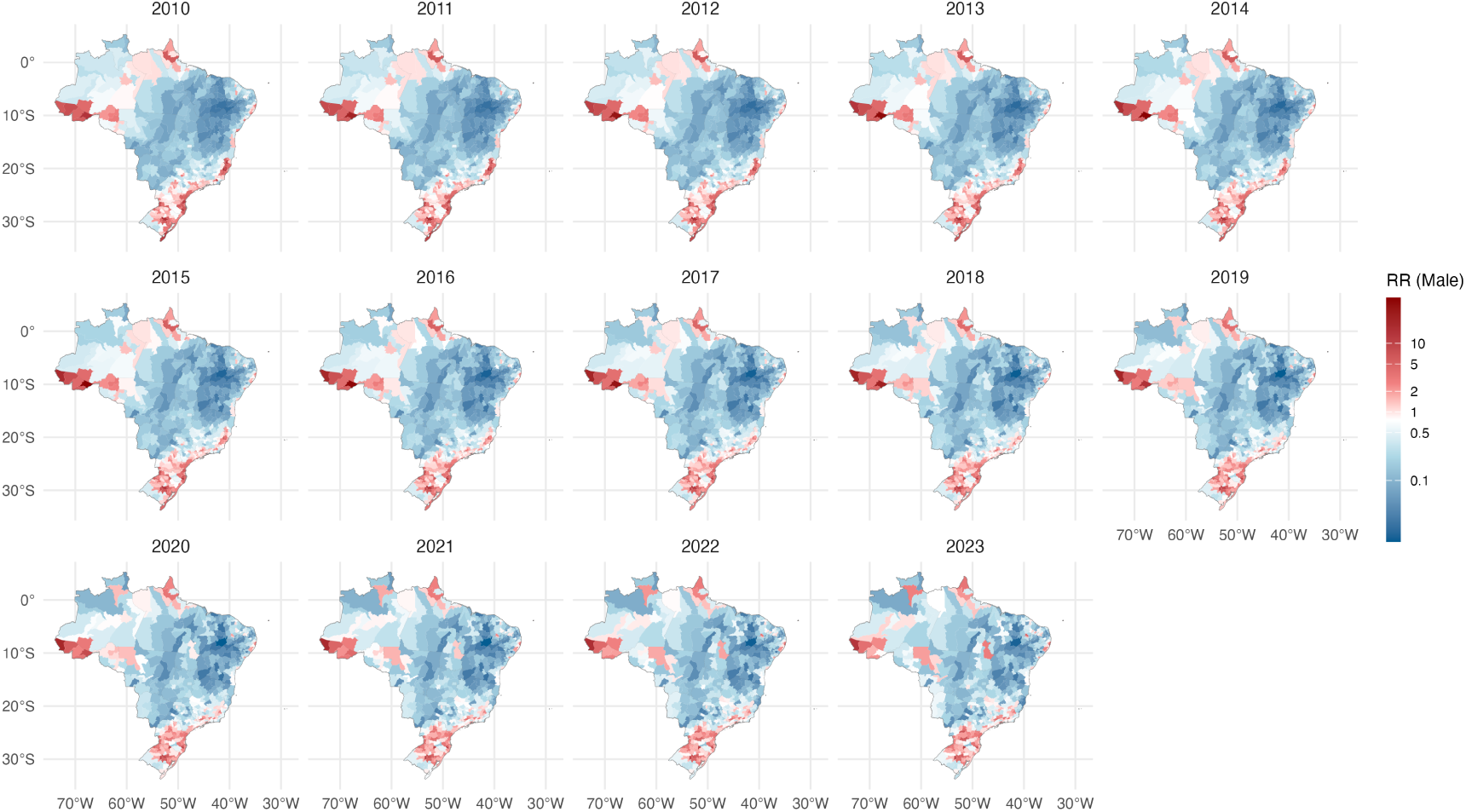
Relative risk (male) of leptospirosis in Brazil (2010–2023). Microregion-level estimates (*RR >* 1 indicates elevated risk)

**Figure 7:**
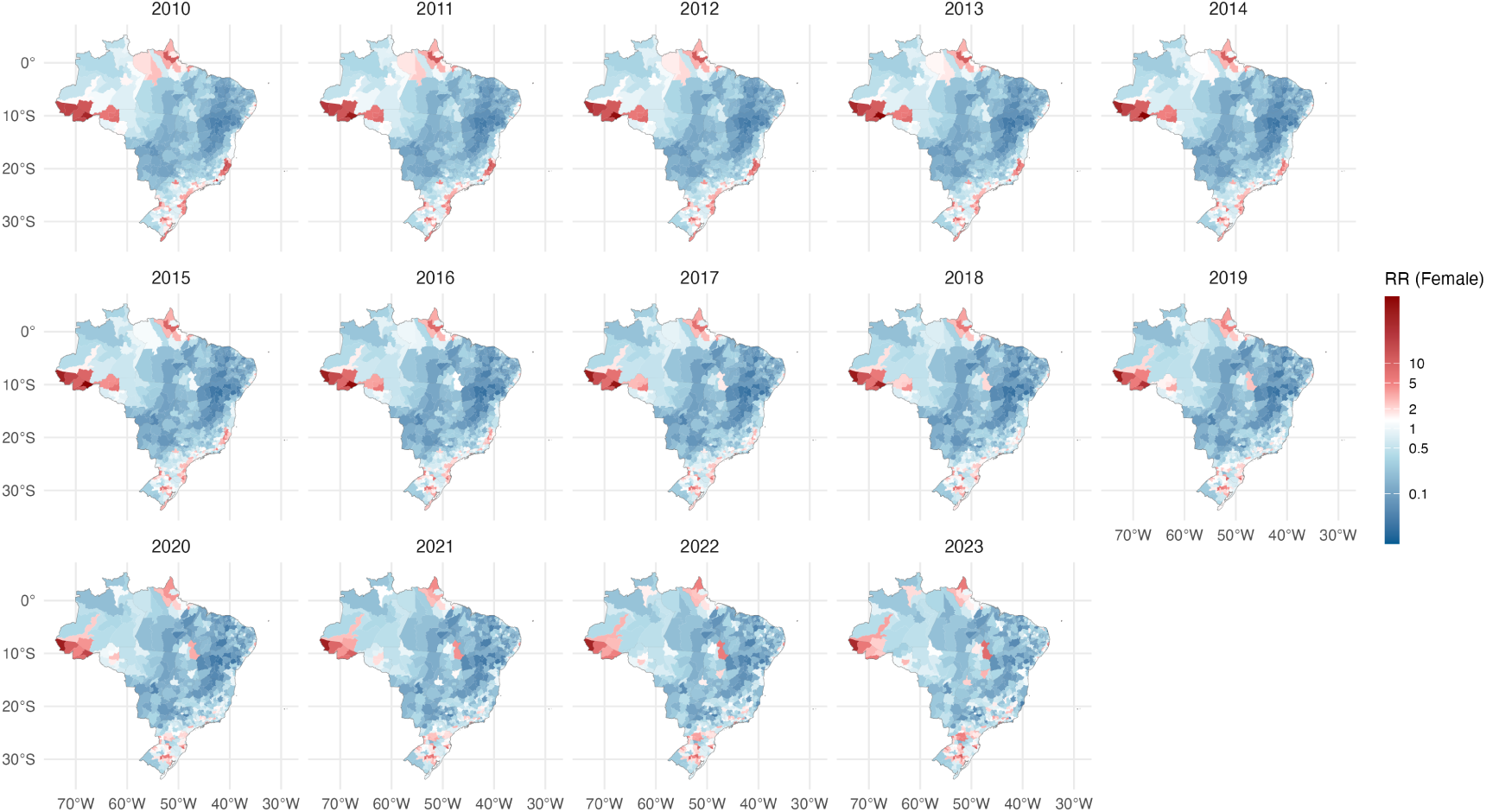
Relative risk (female) of leptospirosis in Brazil (2010–2023). Microregion-level estimates (*RR >* 1 indicates elevated risk)

**Figure 8:**
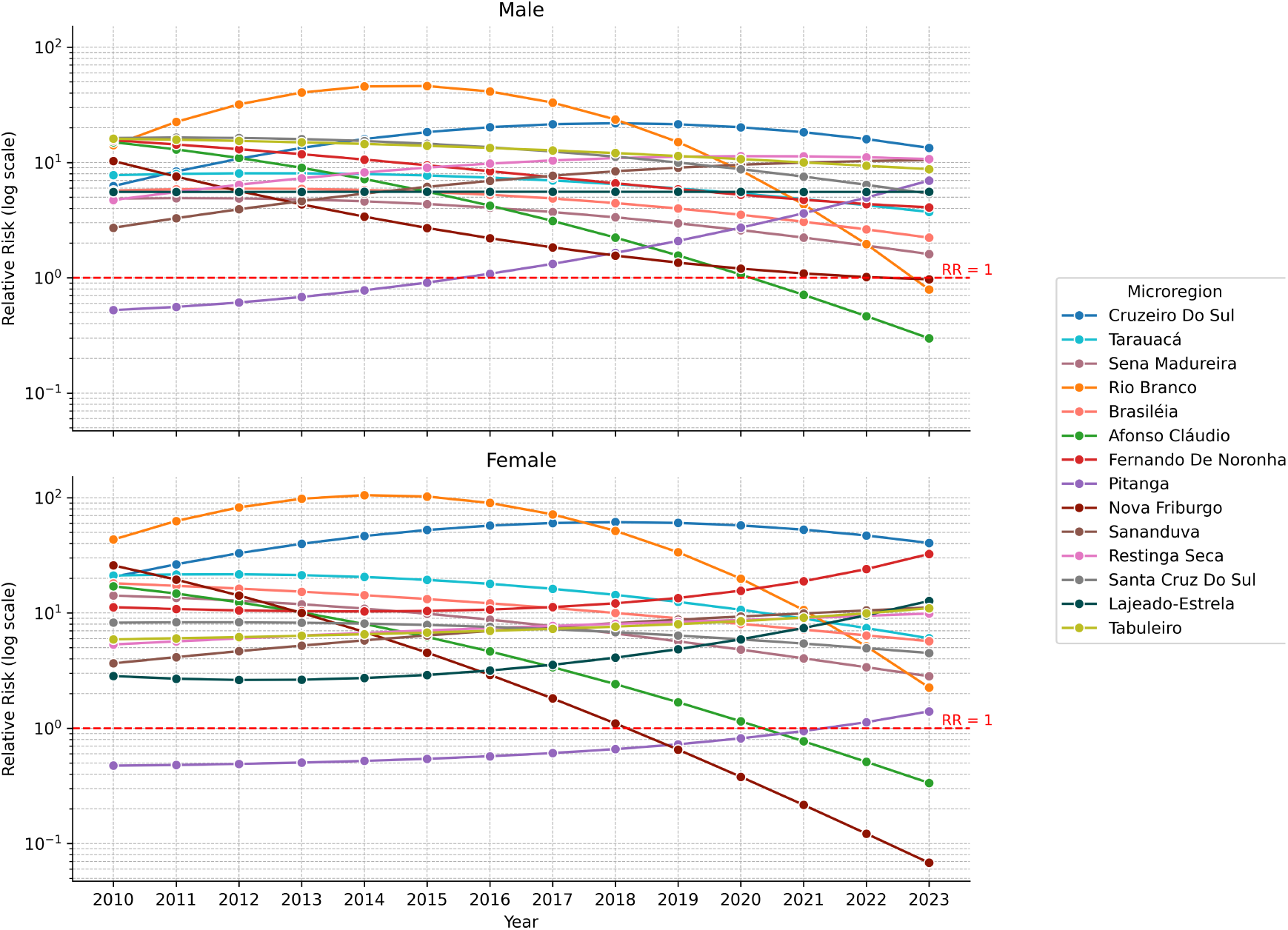
Relative risk (male and female) Over Time for top microregions from 2010 to 2023.

For males, several regions showed consistently elevated risk over time. Notably, parts of Acre and Rondônia presented years with male *RR* values exceeding 20, indicating persistent elevated risk within the male population. Similarly, multiple microregions in southern Brazil—particularly in Santa Catarina and Rio Grande do Sul—showed male *RR* values greater than 5 across several years. Select coastal and riverine microregions, such as Fernando de Noronha (PE), where male *RR* peaked sharply in early years, reflecting an intense but localized surge in incidence.

The female *RR* map also revealed patterns of elevated risk, though these were more temporally intermittent and geographically constrained. In particular, Rio Branco (AC) experienced substantial increases in female *RR* during outbreak years, with values exceeding 10. Elevated female *RR* was also noted sporadically in Espírito Santo, Pernambuco, and several microregions in the South.

Despite the separate modeling, these maps provide critical insights into vulnerable populations that may be obscured in aggregated analyses. The identification of female-specific hotspots underscores the need for gender-sensitive surveillance and intervention strategies, particularly in regions where access to care, health-seeking behavior, or ex-posure pathways may differ by gender.

Together, these gender-specific *RR* maps emphasize the importance of targeted interventions, such as occupational risk mitigation and gender-aware education cam-paigns, especially in the Amazon Basin and southern Brazil where long-term endemicity continues to disproportionately affect male populations.

To further investigate extreme cases, we selected the top 5 microregions with the highest *RR* for either males or females in any given year between 2010 and 2023 (Fig-ure 8). While some regions exhibited similar temporal trajectories for both genders, others revealed striking gender disparities in risk evolution.

The most prominent high-risk areas were the microregions of Rio Branco (AC) and Cruzeiro do Sul (AC), which consistently ranked among the top for both genders. In Rio Branco, male *RR* peaked in 2015 at 46.01 (95% CI: 43.55–48.57), while female *RR* reached an even higher peak in 2014 at 105.26 (95% CI: 98.12–112.78). Both genders exhibited a sharp decline in *RR* after these peak years. Similarly, in Cruzeiro do Sul, risk rose steadily until 2018, with male *RR* peaking at 21.89 (95% CI: 19.57–24.42) and female *RR* at 61.43 (95% CI: 53.58–70.12), followed by a subsequent downward trend.

In contrast, certain microregions demonstrated clear gender divergence. In Tabuleiro (SC), male *RR* declined steadily over time, while female *RR* increased, suggesting potential gender-specific exposure patterns or healthcare access differences. A similar divergence was observed in Lajeado-Estrela (RS), where male *RR* remained relatively stable, whereas female *RR* gradually increased throughout the study period.

Nova Friburgo (RJ) illustrates a case of gender-converging low risk. The male *RR* steadily declined toward the baseline of 1.0, reaching 0.97 (95% CI: 0.51–1.68) in 2023. Female *RR* dropped even more sharply, falling from 1.10 (95% CI: 0.52–2.06) in 2018 to 0.07 (95% CI: 0.008–0.26) in 2023, indicating a substantial reduction in disease burden for women in that region.

Finally, Pitanga (PR) presents a case of simultaneous increase for both genders, albeit with different timelines. Male *RR* exceeded 1.0 in 2016 (1.08, 95% CI: 0.63–1.75), while female *RR* crossed the threshold only in 2022 (1.13, 95% CI: 0.10–4.79), suggesting a delayed but emerging risk among women.

### 3.5 Interactive web application

In addition to the static visualizations, we developed an interactive web dashboard to facilitate exploration of the Bayesian spatio-temporal model estimates: https://paulamoraga.shinyapps.io/leptobrazilapp/. The application allows users to navigate spatio-temporal patterns of leptospirosis across Brazil’s microregions from 2010 to 2023. Users can interactively view maps of estimated relative risk (*RR*), population size, and reported cases, disaggregated by gender and year. The dashboard also includes tabular summaries and time-series visualizations, offering an intuitive platform to explore model outputs and epidemiological trends. This tool is intended to support researchers and public health officials in identifying priority areas and designing targeted, data-driven interventions.

## 4 Discussion

This study provides a comprehensive spatio-temporal analysis of leptospirosis in Brazil from 2010 to 2023, leveraging national surveillance data and advanced statistical methods to characterize disease burden and risk across microregions and demographic groups. Our findings underscore the persistent spatial heterogeneity of leptospirosis in Brazil and highlight the importance of disaggregated, locally tailored public health responses.

The descriptive analysis confirmed leptospirosis as a substantial public health concern, with nearly 50,000 reported cases during the study period. The burden was disproportionately concentrated among males (80.1%), consistent with established literature attributing higher risk to occupational exposure, particularly in agriculture, waste management, and flood-prone settings [29, 30, 31, 32]. Temporally, the national case count fluctuated markedly, with prominent peaks in 2011, 2013, and 2018. A sharp decline in 2020 and 2021 coincided with the COVID-19 pandemic, possibly due to reduced exposure or underreporting. Case counts began to rise again post-pandemic, indicating a return to baseline transmission conditions.

Spatio-temporal cluster analysis using SaTScan revealed 30 statistically significant high-risk clusters. These clusters were predominantly located in the southern states of Rio Grande do Sul, Santa Catarina, and Parańa, as well as in western and northern states including Acre and Rondônia. This spatial pattern aligns with previous studies that emphasize environmental and ecological conditions—such as high rainfall, urban flooding, and rodent proliferation—as key drivers of transmission [33, 34]. An interactive web-based map was developed to visualize these clusters, offering public health authorities a tool to explore outbreak areas and prioritize interventions geographically.

Bayesian modeling using INLA produced relative risk (*RR*) estimates for all 558 microregions annually. Microregions in southern Brazil and the western Amazon consistently showed *RR* values well above 1, with some exceeding 20 in specific years. The temporal *RR* trends also reflected national patterns, with elevated risk in peak outbreak years and substantial reductions during the COVID period. A derived map summarizing the number of years each microregion experienced *RR >* 1 highlighted areas of persistent endemicity, particularly in Acre, Rondônia, and the South.

A critical addition to this analysis was the disaggregation of risk by gender. Gender-specific models revealed distinct gender-based risk trajectories. For instance, Rio Branco and Cruzeiro do Sul exhibited high *RR* for both genders, peaking around 2014–2018 before declining. By contrast, microregions like Tabuleiro (SC) and Lajeado-Estrela (RS) showed diverging trends, with male risk stable or declining while female risk increased. In Nova Friburgo (RJ), *RR* declined for both genders, but more sharply among females. Pitanga (PR) showed increasing risk over time in both genders, but with delayed elevation among females. These patterns highlight not only shared exposure settings but also gender-specific vulnerabilities and temporal lags in risk emergence.

Methodologically, this study contributes to the growing body of work integrating spatio-temporal disease mapping with Bayesian modeling in Latin America [35, 36]. The use of INLA offers a computationally efficient alternative to traditional MCMC approaches, allowing for estimation over a large number of spatial units and time points. Our incorporation of both SaTScan and INLA provided complementary in-sights—detecting outbreaks via cluster detection while capturing risk variation through Bayesian inference.

Several limitations warrant consideration. First, the accuracy of SINAN data may be affected by underreporting, particularly during overlapping public health crises such as COVID-19. Second, while our analysis accounted for space and time, it did not incorporate environmental or climatic covariates, which could enhance risk modeling [37, 38, 39, 40]. Lastly, data was disaggregated by gender but there was not information for other strata, precluding analyses of other vulnerable subgroups.

Despite these limitations, our findings provide robust, data-driven evidence to in-form leptospirosis surveillance and control across Brazil. The identification of persistent high-risk microregions—particularly in Acre, Rondônia, and throughout the southern states—highlights the urgent need for sustained, locally targeted public health inter-ventions. These regions often face overlapping structural vulnerabilities, including in-adequate sanitation, recurrent flooding, and limited access to timely healthcare, all of which may contribute to persistent endemic transmission.

The clear gender disparities observed in our analysis underscore the importance of integrating gender-disaggregated surveillance and response mechanisms into national health systems [41]. While in some regions males exhibited consistently higher risk, fe-male patterns revealed delayed or emerging risk, suggesting overlooked vulnerabilities in exposure pathways, care access, or awareness. Addressing these disparities will require not only gender-sensitive interventions but also broader improvements in infrastructure, risk communication, and social protection.

By combining national-scale surveillance data with spatio-temporal modeling and interactive tools, this study contributes both methodologically and practically to ef-forts aimed at strengthening regional health surveillance and response. Future research should incorporate additional environmental, occupational, and socioeconomic deter-minants to enhance predictive capacity and intervention targeting. Ultimately, ad-vancing equity-oriented, precision public health approaches will be critical to reducing the burden of leptospirosis and other climate-sensitive diseases across Brazil and Latin America.

## 5 Conclusion

This study provides the first nationwide, microregion-level spatio-temporal analysis of leptospirosis in Brazil spanning over a decade. By integrating national surveillance data with Bayesian modeling and spatial cluster detection, we identified persistent high-risk areas, shifting transmission dynamics, and marked gender disparities in risk. The results emphasize the spatial heterogeneity of leptospirosis burden across Brazil and highlight the need for localized, data-informed strategies.

Our findings underscore the importance of routine, disaggregated disease surveil-lance and the utility of interactive tools for enhancing epidemiological insight and pub-lic health planning. Continued investment in targeted interventions, particularly in historically under-resourced and environmentally vulnerable regions, will be essential to reduce leptospirosis burden and address structural determinants of exposure. As climate variability, urbanization, and social inequities continue to shape disease risk, precision public health approaches must be prioritized to ensure equitable and effective responses.

## Data Availability

All data used in this study is open access and described in the "Material and methods" section.

## Declarations

### Ethical approval and consent to participate

Not applicable.

### Consent for publication

Not applicable.

### Availability of data and materials

All data used in this study is open access and described in the “Material and methods” section. Additionally, an interactive web application for further exploration and visualization of the risk estimates is available at https://paulamoraga.shinyapps.io/leptobrazilapp/. The cluster map can be accessed at https://www.paulamoraga.com/leptobrazilclusters/.

### Authors’ contributions

PM and XC conceived and designed the study. XC collected and processed the data. PM and XC performed the experiments. All the figures and maps are drawn by XC. XC and PM were contributors in writing and revising the manuscript. All authors read and approved the final manuscript.

## Acknowledgements

Not applicable.

## References

[1] Costa F, Hagan JE, Calcagno J, Kane M, Torgerson P, Martinez-Silveira MS, et al. Global morbidity and mortality of leptospirosis: a systematic review. PLoS neglected tropical diseases. 2015;9(9):e0003898.

[2] Hagan JE, Moraga P, Costa F, Capian N, Ribeiro GS, Wunder Jr EA, et al. Spatiotemporal determinants of urban leptospirosis transmission: four-year prospective cohort study of slum residents in Brazil. PLoS neglected tropical diseases. 2016;10(1):e0004275.

[3] Ko AI, Goarant C, Picardeau M. Leptospira: the dawn of the molecular genetics era for an emerging zoonotic pathogen. Nature Reviews Microbiology. 2009;7(10):736–47.

[4] Bharti AR, Nally JE, Ricaldi JN, Matthias MA, Diaz MM, Lovett MA, et al. Leptospirosis: a zoonotic disease of global importance. The Lancet infectious diseases. 2003;3(12):757–71.

[5] Riley LW, Ko AI, Unger A, Reis MG. Slum health: diseases of neglected populations. BMC international health and human rights. 2007;7:1–6.

[6] Reis RB, Ribeiro GS, Felzemburgh RD, Santana FS, Mohr S, Melendez AX, et al. Impact of environment and social gradient on Leptospira infection in urban slums. PLoS neglected tropical diseases. 2008;2(4):e228.

[7] Schneider MC, Leonel DG, Hamrick PN, Caldas EPd, Veĺasquez RT, Paez FAM, et al. Leptospirosis in Latin America: exploring the first set of regional data. Revista Panamericana de Salud Pública. 2018;41:e81.

[8] Matsushita N, Ng CFS, Kim Y, Suzuki M, Saito N, Ariyoshi K, et al. The non-linear and lagged short-term relationship between rainfall and leptospirosis and the intermediate role of floods in the Philippines. PLoS neglected tropical diseases. 2018;12(4):e0006331.

[9] Radi MFM, Hashim JH, Jaafar MH, Hod R, Ahmad N, Nawi AM, et al. Leptospirosis outbreak after the 2014 major flooding event in Kelantan, Malaysia: a spatial-temporal analysis. The American journal of tropical medicine and hygiene. 2018;98(5):1281.

[10] López MS, Müller GV, Lovino MA, Gómez AA, Sione WF, Pomares LA. Spatio-temporal analysis of leptospirosis incidence and its relationship with hydroclimatic indicators in northeastern Argentina. Science of the Total Environment. 2019;694:133651.

[11] Baquero OS, Machado G. Spatiotemporal dynamics and risk factors for human Leptospirosis in Brazil. Scientific reports. 2018;8(1):15170.

[12] Galan DI, Roess AA, Pereira SVC, Schneider MC. Epidemiology of human leptospirosis in urban and rural areas of Brazil, 2000–2015. PloS one. 2021;16(3):e0247763.

[13] Melchior LAK, da Silva KRC, Silva AEP, Chiaravalloti-Neto F. Ańalise espacial, tem-poral e espaçotemporal dos casos de leptospirose no Acre, 2001-2022. Revista Brasileira de Epidemiologia. 2024;27:e240063.

[14] Melchior LAK, da Silva KRC, Silva AEP, Chiaravalloti-Neto F. Spatial, temporal, and space-time analysis of leptospirosis cases in Acre, 2001-2022. Revista Brasileira de Epidemiologia. 2024;27:e240063.

[15] Instituto Brasileiro de Geografia e Estatística (IBGE). IBGE: Brazil’s population reaches 212.6 million; 2024. https://www.gov.br/secom/en/latest-news/2024/08/ibge-brazils-population-reaches-212-6-million.

[16] Ministério da Saúde, Brasil. Sistema de Informação de Agravos de Notificação (SINAN); 2023. Available at: http://www.saude.gov.br/sinan.

[17] Ministério da Saúde. População Residente -Estudo de Estimativas Populacionais por Munićıpio, Idade e Sexo 2000-2024 -Brasil; 2024. DATASUS -Tecnologia da Informação a Serviço do SUS, Informações de Saúde. http://tabnet.datasus.gov.br/cgi/deftohtm.exe?ibge/cnv/popsvs2024br.def.

[18] Kulldorff M, Inc IMS. SaTScan™: Software for the spatial and space-time scan statistics; 2024. SaTScan™ is a trademark of Martin Kulldorff. https://www.satscan.org.

[19] Kulldorff M. A spatial scan statistic. Communications in Statistics-Theory and methods. 1997;26(6):1481–96.

[20] Rubuga FK, Moraga P, Ahmed A, Siddig E, Remera E, Moirano G, et al. Spatio-temporal dynamics of malaria in Rwanda between 2012 and 2022: a demography-specific analysis. Infectious Diseases of Poverty. 2024;13(1):67.

[21] Moraga P, Kuldorff M. Detection of spatial variations in temporal trends with a quadratic function. Statistical Methods in Medical Research. 2013;25(4):1422–37.

[22] Moraga P. Geospatial health data: Modeling and visualization with R-INLA and shiny. Chapman and Hall/CRC; 2019.

[23] Rue H, Martino S, Chopin N. Approximate Bayesian inference for latent Gaussian models by using integrated nested Laplace approximations. Journal of the Royal Statistical Society Series B: Statistical Methodology. 2009;71(2):319–92.

[24] Moraga P. Small area disease risk estimation and visualization using R. The R Journal. 2018;10(1):495–506.

[25] Besag J, York J, Mollíe A. Bayesian image restoration, with two applications in spatial statistics. Annals of the institute of statistical mathematics. 1991;43:1–20.

[26] Haake DA, Levett PN. Leptospirosis in humans. Leptospira and leptospirosis. 2014:65–97.

[27] Luiz-Silva W, Oscar-Júnior AC. Climate extremes related with rainfall in the State of Rio de Janeiro, Brazil: a review of climatological characteristics and recorded trends. Natural Hazards. 2022;114(1):713-32.

[28] Alves PJ, de Andrade Lima RC, Emanuel L. Natural disasters and establishment performance: Evidence from the 2011 Rio de Janeiro Landslides. Regional Science and Urban Economics. 2022;95:103761.

[29] Hartskeerl R, Collares-Pereira M, Ellis W. Emergence, control and re-emerging leptospirosis: dynamics of infection in the changing world. Clinical microbiology and infection. 2011;17(4):494–501.

[30] Londe LdR, Da Conceição RS, Bernardes T, Dias MCdA. Flood-related leptospirosis outbreaks in Brazil: perspectives for a joint monitoring by health services and disaster monitoring centers. Natural Hazards. 2016;84(2):1419–35.

[31] Brakenridge GR. Global Active Archive of Large Flood Events; 2016. Dartmouth Flood Observatory, University of Colorado. http://floodobservatory.colorado.edu/Archives/.

[32] Global Disaster Alert and Coordination System (GDACS). GDACS: Global Disaster Alert and Coordination System;. European Commission and United Nations. https://www.gdacs.org/.

[33] Teles AJ, Bohm BC, Silva SCM, Bruhn FRP. Socio-geographical factors and vulnerability to leptospirosis in South Brazil. BMC public health. 2023;23(1):1311.

[34] Teles AJ, Bohm BC, Silva SCM, Bruhn NCP, Bruhn FRP. Spatial and temporal dynamics of leptospirosis in South Brazil: A forecasting and nonlinear regression analysis. PLoS neglected tropical diseases. 2023;17(4):e0011239.

[35] Ortega-Lenis D, Arango-Londoño D, Herńandez F, Moraga P. Effects of climate variability on the spatio-temporal distribution of Dengue in Valle del Cauca, Colombia, from 2001 to 2019. PLoS One. 2024;19(10):e0311607.

[36] Gómez MJ, Barboza LA, Moraga PVP. Bayesian spatial modeling of childhood over-weight and obesity prevalence in Costa Rica. BMC Public Health. 2023;23(651).

[37] Llop MJ, Gómez A, Llop P, Ĺopez MS, Müller GV. Prediction of leptospirosis outbreaks by hydroclimatic covariates: a comparative study of statistical models. International Journal of Biometeorology. 2022;66(12):2529–40.

[38] Chen X, Moraga P. Assessing dengue forecasting methods: a comparative study of statistical models and machine learning techniques in Rio de Janeiro, Brazil. Tropical medicine and health. 2025;53(1):52.

[39] Chen X, Moraga P. Forecasting dengue across Brazil with LSTM neural networks and SHAP-driven lagged climate and spatial effects. BMC public health. 2025;25(1):973.

[40] Marteli AN, Guasselli LA, Diament D, Wink GO, Vasconcelos VV, et al. Spatio-temporal analysis of leptospirosis in Brazil and its relationship with flooding. Geospatial Health. 2022;17(2).

[41] World Health Organization and others. Addressing sex and gender in epidemic-prone infectious diseases. In: Addressing sex and gender in epidemic-prone infectious diseases; 2007.

